# A study of PROGRESS: the Therapeutic Potential of 17 OHPC on the Pathophysiology of Severe Preeclampsia

**DOI:** 10.64898/2026.07.15.26358196

**Authors:** Charles Brewerton, Christy L. Chambers, Sheila Belk, Kedra Wallace, Montianah Roseburg, Nathan Campbell, Yelena R. Neely, Cameronne A. Dodd, Rachael Morris, Sarah Novotny, James M. Tucker, Babbette LaMarca, Lorena M. Amaral

**Affiliations:** Obstetrics and Gynecology, University of Mississippi Medical Center, Jackson, MS, 39216; Pharmacology & Toxicology, University of Mississippi Medical Center, Jackson, MS, 39216

**Author notes:** **Address correspondence to:** Lorena M. Amaral, MS, PhD, FAHA, Associate Professor, Department of Pharmacology & Toxicology, University of Mississippi Medical Center, 2500 North State Street, Jackson, Mississippi 39216.

**Keywords:** Preeclampsia, pregnancy, blood pressure, progesterone

## Abstract

Preeclampsia (PE), new onset hypertension after 20 weeks of gestation, affects 10% of all pregnancies in the U.S. and it is associated with progesterone deficiency, chronic inflammation, elevated angiotensin II type 1 receptor agonistic autoantibody (AT1-AA) and endothelial dysfunction. Progesterone, through its receptors, stimulates an anti-inflammatory protein called <u>P</u>rogesterone Induced <u>B</u>locking <u>F</u>actor (PIBF) which decreases during various pregnancy disorders. Therefore, this study was designed to test the hypothesis that a progestogen, in the form of 17-hydroxyprogesterone caproate, stimulates PIBF, lowers vasoactive mechanisms which reduces maternal blood pressure in women with early-onset preeclampsia (EOPE). PE women received 17-OHPC (250 mg, I.M.) and blood draws were collected before and after 17-OHPC supplementation. Placentas were collected at the delivery. 17-OHPC prolonged time of delivery beyond 72h on average and maternal blood pressure was significantly decreased in PE+17-OHPC. Progesterone and PIBF levels were reduced in PE group vs. NP group. Importantly, 17-OHPC increased PIBF and decreased vasoactive mechanisms and markers of inflammation. In conclusion, 17-OHPC or progesterone supplementation improves maternal outcomes in response to EOPE without causing further harm to the fetus.

## 1. INTRODUCTION

Preeclampsia (PE) is a multisystem hypertensive disorder occurring in the second half of pregnancy and postpartum associated with proteinuria, severe blood pressure elevation, and end organ damage. PE affects 10% of all pregnancies in the U.S. and occurs with markedly increased frequency with advancing maternal age and in black and non-white ethnic groups[1, 2]. In the US, both the incidence and prevalence of PE have been increasing[3, 4]. Hypertensive disorders of pregnancy complicate up to 20.9% of delivery hospitalizations in the US in persons identifying as African American, 31% in persons over 45 years of age, and 31.6% of patients whose delivery admission resulted in a maternal death [1, 2]. Despite being a leading cause of maternal, fetal, and neonatal morbidity and mortality, timely delivery of the pregnancy is currently the best known treatment to manage the end organ damage associated with preeclampsia[4].

PE has been subcategorized by gestational age at symptom onset. Early-onset preeclampsia (EOPE) occurs prior to 34 weeks of gestation and PE after 34 weeks of gestation is termed late-onset preeclampsia (LOPE)[5, 6]. EOPE tends to be a more severe disease phenotype with worse maternal and fetal outcomes[7]. PE represents a dichotomy of risk where the benefits of immediate delivery must be weighed against continuation of pregnancy to reduce prematurity. The American College of Obstetricians and Gynecologists (ACOG) oppose expectant management of PE with severe features after 34 weeks of gestation due to associated health risks[4]. However, the risks of prematurity prior to 34 weeks of gestation are great enough to consider expectant management in the absence of contraindications. Our study is focused on modifying outcomes for EOPE patients who are candidates for expectant management.

The entire pathophysiology of PE has not been fully elucidated and is known to be multifactorial. Placental endovascular dysfunction and immune dysregulation have been well studied as hallmarks of PE. The endocrine pathway through which progesterone has its immune modulatory effects is not yet fully understood. The major sources of progesterone in the female reproductive life cycle are the corpus luteum and the placenta after the first 8-10 weeks of pregnancy[8]. Progesterone is known to stimulate the production of progesterone-induced blocking factor (PIBF). PIBF is secreted by progesterone-exposed lymphocytes. It is a soluble endocrine protein that exhibits a downstream anti-inflammatory effect on natural killer cells (NKC) and T cells by modulating cytokine production and shifting T-cell populations from CD4+TH1 to CD4+TH2 predominance[9]. Treg and Th2 predominance favors immune tolerance of the pregnancy and is associated with normal pregnancy outcomes, where as NKC and CD4+ T cell activation is associated with miscarriage and PE.

As the number of CD4+TH1 cells are increased, the release of cytokines and B cells producing autoantibodies (AT1-AA) that activate the renin angiotensin system are increased. These changes also cause increased expression of vasoconstrictors Endothelin-1 (ET-1), soluble vascular endothelial growth factor receptor 1 (sFlt-1), and soluble endoglin (sEng)[10–13]. Conversely, vasodilators like nitric oxide (NO) are decreased in PE. These immune and vascular dysfunctions work cyclically by inducing a progressively more robust inflammatory and vasoconstrictor and hypertensive response. [14]. Thus the clinical manifestations of this pathway are hypertension, endothelial dysfunction, placental insufficiency, and fetal growth restriction (FGR) in association with chronic inflammation[15].

We have shown that, in our patient population, patients with PE have lower levels of circulating progesterone[16]. Progesterone deficiency in the setting of PE is established in the literature and therefore a PIBF deficiency is reasonably hypothesized. Although, this has not yet been demonstrated in a human study[17–20]. Our pre-clinical data shows that 17 OHPC improve the pathology of placental ischemia in rats models of PE and that PIBF administration to rat models of PE attenuates the hypertensive response to both pro-inflammatory mediators and the reduced uterine perfusion procedure[9, 21, 22] Additionally, we have previously shown that PIBF blockade causes hypertension in control pregnant rats[23].

Treatment of preeclampsia with progesterone has had limited evaluation, but results have been promising. Sammour et al. published in 1975 that 80% of 40 PE patients after 34 weeks of gestation had a significant decrease in blood pressure and increased urine output following treatment with IM progesterone[17]. In 1982, Sammour et al. reported improved blood pressure and urine output in a second cohort of patients with severe pregnancy-related hypertension treated with progesterone[24]. It is worth noting in these two trials that the peak antihypertensive effect of progesterone was noted between six and twelve days. In 2023, Liu et al. demonstrated an improvement in preeclampsia in patients received oral micronized progesterone[20]. Beyone pregnancy, progesterone dosing has also been shown to reduce hypertension in men and postmenopausal females.

The present study focuses on treatment of PE, though progestogens have also been evaluated for prevention of PE. A 2006 Cochrane review concluded that there was insufficient evidence to support the use of progestogens for the prevention of preeclampsia based on four studies with variable doses and routes of either 17-OHPC or micronized progesterone[25]. While Aspirin currently remains the only recommended medication for prevention of PE, a subsequent metanalysis by Melo et al. in 2024 concluded that vaginal micronized progesterone was effective in prevention of PE if initiated in the first trimester[26].

Therefore, given the promising clinical and preclinical data regarding progestogens and its effects on hypertension and PE, we hypethosized that 17-OHPC might allow patients with EOPE with severe features to safely extend their pregnancy for fetal benefit. We hypothesized that administration of 17-OHPC to persons with EOPE with severe features would result in decreased maternal blood pressures, decreases in biomarkers of preeclampsia, increased circulating PIBF, and increased latency to delivery due to amelioration of the disease state. Our research represents, to our knowledge, the first time that 17-OHPC administration has been studied for treatment rather than prevention of preeclampsia.

## 2. MATERIAL AND METHODS

### Ethics Statement and inclusion criteria

The protocol was approved by the Ethics Committee on Human Investigation at the University of Mississippi Medical Center (UMMC) (Protocol IRB number: 2015-0029). Patients were admitted to Winfred Wiser Hospital for Women and Infants (Jackson, MS) with evidence of severe PE. Pregnant women meeting ACOG criteria for PE during pregnancy[27] were evaluated by co-a maternal and fetal medicine specialist in OB/GYN at UMMC. The treating physician immediately contacted the MFM fellow on call and the Research Division personnel to facilitate the initiation of study procedures, including the consent process and blood draws. UMMC antepartum patients between the ages of 18-45 with preterm PE between 23w0 and 34w0d gestation when initially evaluated without fetal anomaly or maternal compromise requiring delivery, not already receiving 17-OHPC for other reason, and were able to understand English were included in the study. Patients consented to have clinical data, blood and placenta collected. Blood and placentas were obtained from 18 PE treated with 17-OHPC, 17 PE untreated with 17-OHPC, and 8 normotensive pregnant women. Typical expectant management of EOPE with severe features at our institution includes inpatient admission with close and frequent maternal and fetal monitoring, steroids to promote fetal maturation, interval use of magnesium sulfate to reduce risk of eclamptic seizures, and use of antihypertensive medications such as labetalol, hydralazine, or nifedipine both scheduled and as needed. All EOPE patients with severe features received this standard of care treatment with or without 17-OHPC administration.

Inclusion criteria were maternal age 18-45 years old, gestational age between 23w0d and 34w0d, willing and able to understand and consent to study participation. Patients were excluded from enrollment in the setting of severe fetal anomalies, patients already taking 17-OHPC for another indication, maternal compromise requiring emergent delivery (i.e. ongoing placental abruption, DIC, pulmonary edema), fetal compromise requiring emergent delivery (i.e. fetal bradycardia, recurrent late fetal heart rate decelerations, minimal to absent fetal heart rate variability, biophysical profile less than 6/10), and any clinical condition that was thought to reasonably exclude a patient from expectant management of EOPE including those listed in ACOG practice guidelines (fetal demise, persistent reversed end diastolic flow, eclampsia, HELLP) and conditions that were not routinely managed expectantly at our institution at the time of study conception including platelet count of less than 100,000 per microliter, elevated transaminases double the upper limit of normal, severe oligohydramnios (AFI>5), and fetal growth restriction less than the 5^th^ percentile. Of note, while our institutional practice no longer considers the foregoing measures of platelet count, serum transaminases, restricted fetal growth, or amniotic fluid volume to represent an automatic exclusion to expectant management, these exclusion criteria were retained to maintain uniform enrollment practices over the study period.

### Enrollment Maternal and fetal outcomes

Enrollment began in 2017 and continued into 2026. Patients were enrolled by our Pharmacology Clinical Research Core after confirmation of the diagnosis of preeclampsia with severe features and review of inclusion and exclusion criteria by a maternal fetal medicine fellow. The nurse coordinator participated in 17-OHPC administration, blood collection, and assisted collection of the placenta post-delivery (**Figure 1**). Otherwise, clinical information was collected by research staff. PE treated group received 17-OHPC, 250 mg/mL intramuscular weekly (Marty’s Compounding Pharmacy, Jackson, MS). The dosage and timing of injections mirror what was previously FDA approved for prevention of recurrent preterm birth as it was known to have a good safety profile for pregnancy.

**Figure 1.**
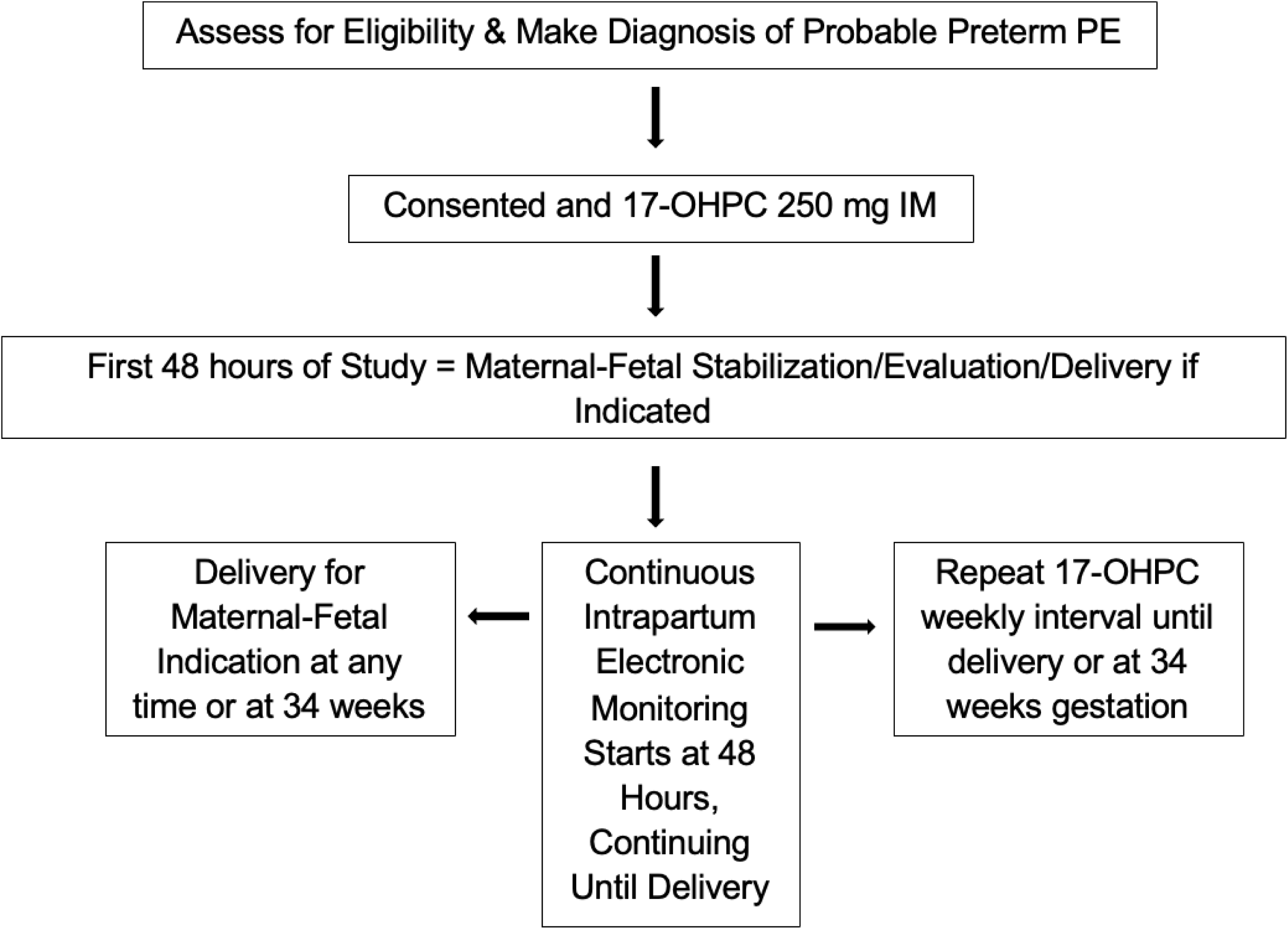
Assess for eligibility and make diagnosis of probable preterm PE.

Primary outcome measured was initially determined to be latency to delivery as a stand in for all cause severity of preeclampsia. Maternal blood pressure over time was assessed at enrollment and daily until delivery. Other secondary outcomes were development of adverse maternal outcomes, adverse fetal or neonatal outcomes, and changes in maternal laboratory values.

Adverse maternal outcomes were defined as placental abruption, pulmonary edema, acute kidney injury, diffuse intravascular coagulopathy, adverse neurologic events (stroke, seizure, and persistent neurologic features including refractory headaches and vision changes), and development of laboratory abnormalities meaning either signs of end organ dysfunction or changes in biomarkers of preeclampsia. Fetal outcomes are measured as decreased, Apgar scores, prolong NICU stay, interventricular hemorrhage, respiratory distress syndrome, necrotizing enterocolitis. Birth weight was also recorded.

### Sample collection

Whole blood was collected in anticoagulant-EDTA treated tubes for plasma separation and without anticoagulant tubes for serum separation at study enrollment, 24 hours after initial 17-OHPC dose, every 72-96 hours during continuing pregnancies, and postpartum. After collection, serum tubes were allowed to clot by leaving it at room temperature, followed by centrifugation at 1,000-2,000 g for 10 minutes. Plasma tubes were centrifugated at 1,000-2,000 g for 10 minutes. The resultant supernatant is designated either plasma or serum and stored at −20°C. Fresh placentas were collected at delivery by the study team and samples were obtained and processed by the clinical core. After study sample collection, placentas were delivered to the clinical pathology department for routine clinical evaluation if ordered by the patients delivering providers.

### Determination of TNF-α, sFlt-1, PIBF, and nitrate-nitrite

Plasma collected from participants groups was measured for TNF-α levels using commercial ELISA kits from R&D Systems (Quantikine, Minneapolis, Minnesota) according to the manufacturer’s instructions. The minimal intra and inter-assay detectable level for TNF-α was 5.1% and 9.7% respectively.

Plasma collected from participants groups were measured for sFlt-1 levels using commercial ELISA kits from R&D Systems (Quantikine, Minneapolis, Minnesota) according to the manufacturer’s instructions. The minimal detectable level for sFlt-1 was intra and inter-assay detectable level for sflt-1 was 2.5% and 6.7%, respectively.

Serum collected from participants groups was measured for PIBF levels using commercial ELISA kits from My BioSource (San Diego, California) according to the manufacturer’s instructions. The minimal intra and inter-assay detectable level for PIBF was 9.0 % and 11.0% respectively.

Plasma nitrate–nitrite (NOx) levels were evaluated using commercial Nitrate– Nitrite Colorimetric Assay Kit from Cayman Chemical (Michigan) following instructions outlined by the manufacturer. The inter-assay coefficient of variation is 3.4%, whereas intra-assay coefficient of variation is 2.7%.

### Determination of AT1-AA production

Serum was analyzed for AT1-AA by cardiomyocyte assay performed in the Pharmacology Clinical Research Core. Antibodies were detected by the chronotropic responses to AT1 receptor mediated stimulation of cultured neonatal rat cardiomyocytes coupled with receptor-specific antagonists as previously described[28, 29] Chronotropic responses were measured and expressed in beats per minute (bpm).

### Determination of placental preproendothelin-1 expression

Real-time polymerase chain reaction (qRT-PCR) was used to determine placental preproendothelin-1 (PPET-1) levels. Placentas were quickly removed from storage at - 80°C and an approximate 30 mg piece of tissue was cut from the whole placenta prior to homogenization in the provided lysis buffer. Total RNA was then extracted from the homogenized tissues using the RNeasy Protect Mini Kit (Qiagen). The isolation procedure was performed according to the manufacturer’s instructions provided. After RNA was isolated, concentration and quality were verified using a spectrophotometer (Eppendorf BioPhotometer). cDNA was synthesized from 1 μg of RNA using the iScript cDNA Synthesis Kit (BioRad). Real-time polymerase chain reaction (qRT-PCR) was performed using iQ SYBR Green Supermix (BioRad) and the CFX96 Touch Real-Time PCR Detection System (BioRad). The following primer sequences provided by Life technologies were used for PPET: forward 1, CTAGGTCTAAGCGATCCTTG, and reverse 1, TCTTTGTCTGCTTGGC. Levels of mRNA were calculated using the mathematical formula 2^−ΔΔCt^ (2^avg. Ct gene of interest − avg Ct beta actin^) recommended by Applied Biosystems (Applied Biosystems User Bulletin, No. 2, 1997).

### Extraction and determination of Circulating Endothelin-1

Extraction of plasma ET-1 was done using Sep-pak Columns (Waters Corporation, Milford, Massachusetts). Briefly each column was activated by washing buffers. Each plasma sample was acidified by adding an equal volume of buffers and centrifuged at 3000g for 15 minutes at 4C. The mixture of plasma and buffer was loaded onto the columns followed by column washing. The absorbed endothelin materials were slowly eluted with buffer into an Eppendorf. The eluent was completely evaporated with a centrifugal evaporator, and the solid was rehydrated with 200 mL of assay buffer (RnD Systems; Minneapolis, Minnesota. Plasma collected from pregnant groups was measured for sFlt-1 levels using commercial ELISA kits from R&D Systems (Quantikine, Minneapolis, Minnesota) according to the manufacturer’s instructions. The ET-1 assay displayed a sensitivity of 13.3 pg/mL, inter-assay variability of 8.9%, and intra-assay variability of 3.4%.

### Determination of progesterone levels

Quantitative determinations of progesterone in serum were performed using a modified cold-induced phase separation (CIPS) extraction protocol. Extracts were separated by liquid chromatography (LC) by reversed phase C18 column microflow liquid chromatography using an Eksigent M5 micro-LC and detected by multiple reaction monitoring (MRM) via electrospray ionization tandem mass spectrometry (MS/MS) with a SCIEX QTrap 7500 triple quadrupole with ion trap. Quantitation of endogenous steroids were performed by internal stable isotope labeled standards (SILS) for each steroidal analyte[30, 31].

### Statistical Analysis

Statistical differences were established using a one-way ANOVA with Holm Sidak multiple comparison post-hoc test (>2 groups) or Student’s *t* test (2 groups). Data are shown as means ± S.E.M. * P< 0.05. * p< 0.05 vs. NP and ^#^p< 0.05 vs. PE.

## 3. RESULTS

### Clinical characteristics of patients and fetal outcomes

Analysis of the clinical characteristics of PE and NT pregnant women (**Table 1**) showed statistical difference in gestational age and blood pressure. Patients with EOPE who were treated with 17-OHPC had a mean gestational age (GA) of 30w0d. Patients with EOPE in the control arm had a mean GA of 30w0d days. Notably normotensive controls had more advanced mean GA at 35w2d. We did not see statistical difference in BMI. As expected, systolic and diastolic blood pressures were significantly higher in the PE group than in the NT group. Importantly, 17-OHPC decreased systolic blood pressure.

**Table 1.** Clinical and laboratory characteristics of pregnant women with pre-eclampsia and normotensive pregnant women. Statistical differences were established using a one-way ANOVA with Holm Sidak multiple comparison post-hoc test. Data are shown as means ± S.E.M. * p< 0.05 *vs.* NT and ^#^p< 0.05 *vs.* PE.

| Characteristics | Normotensive pregnant women (n=8) | Pregnant women with Preeclampsia (n=17) | Pregnant women with Preeclampsia treated with 17-OHPC (n=18) |
| --- | --- | --- | --- |
| Gestational Age (weeks) | 35±2* | 30±2 | 30±2 |
| BMI | 34±3 | 40±3 | 37±2 |
| Systolic Blood Pressure (mmHg) | 117±5* | 147±4 | 136±3# |
| Diastolic Blood Pressure (mmHg) | 73±5* | 89±3 | 81±3 |

We followed adverse maternal and fetal outcomes during the study period and found no cases adverse outcomes associated with the 17-OHPC supplementation.

There were no significant differences in rates of adverse outcomes between the control and treatment arms of the EOPE groups. In our study population, there were no cases of intrauterine fetal demise, intracranial hemorrhage, and necrotizing enterocolitis between the treatment groups. APGARs at 1 and 5 mins were similar between the PE treatment and PE control arms at both 1 and 5 minutes. There was also not a statistically significant difference in birth weight between the PE treatment and PE control groups or difference in the rates of small for gestational age births. There were similar rates of acute respiratory distress syndrome (ARDS) and NICU admission and length of stay in the treatment and control PE groups.

### 17-OHPC lowers systolic blood pressure in pregnant women with PE

Maternal systolic blood pressure was 116 +/−4 mmHg in NP women (n=8), 147 +/− 4 mmHg in PE women (n=17) and reduced to 136 +/− 3 mmHg in PE+17-OHPC women (n=18, p<0.05, **Figure 2**).17-OHPC did not affect the fetal weight, however, due to safer blood pressure control, time to delivery was prolonged beyond 72-96 hours.

**Figure 2.**
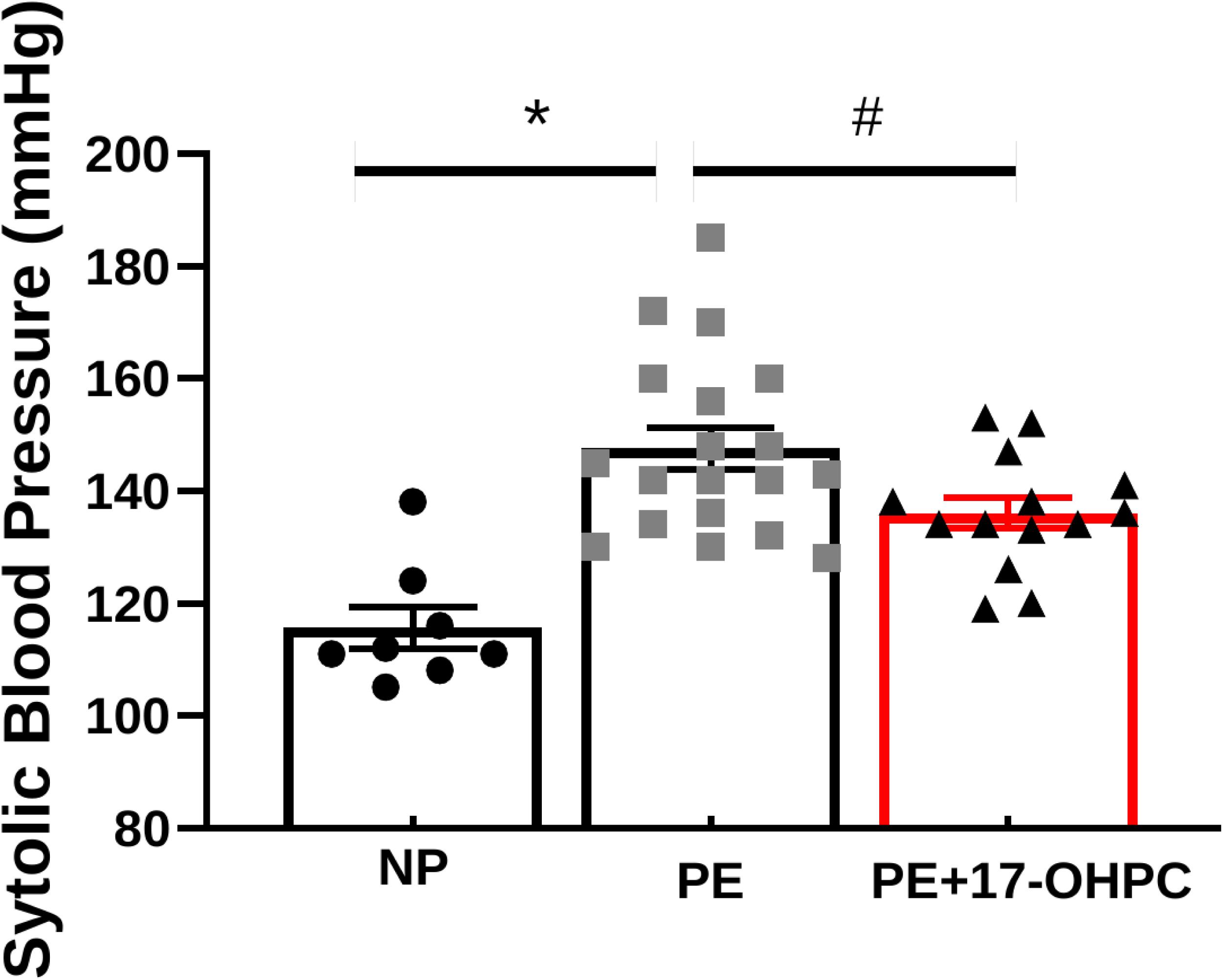
17-OHPC lowers systolic blood pressure in pregnant women with PE. Statistical differences were established using a one-way ANOVA with Holm Sidak multiple comparison post-hoc test. Data are shown as means ± S.E.M. * p< 0.05. * P< 0.05 *vs.* NT and ^#^p< 0.05 *vs.* PE

### Progesterone and PIBF are lower women with PE

Progesterone is reduced in pregnant women with PE compared to normal pregnancy[17–20]. Progesterone levels were 515+/− 65 ng/mL in NP (n=7), 278+/− 46 in PE (n=12 p<0.05), which increased to 391+/−57 ng/mL in PE+17-OHPC group (n=6, **Figure *3***).

**Figure 3.**
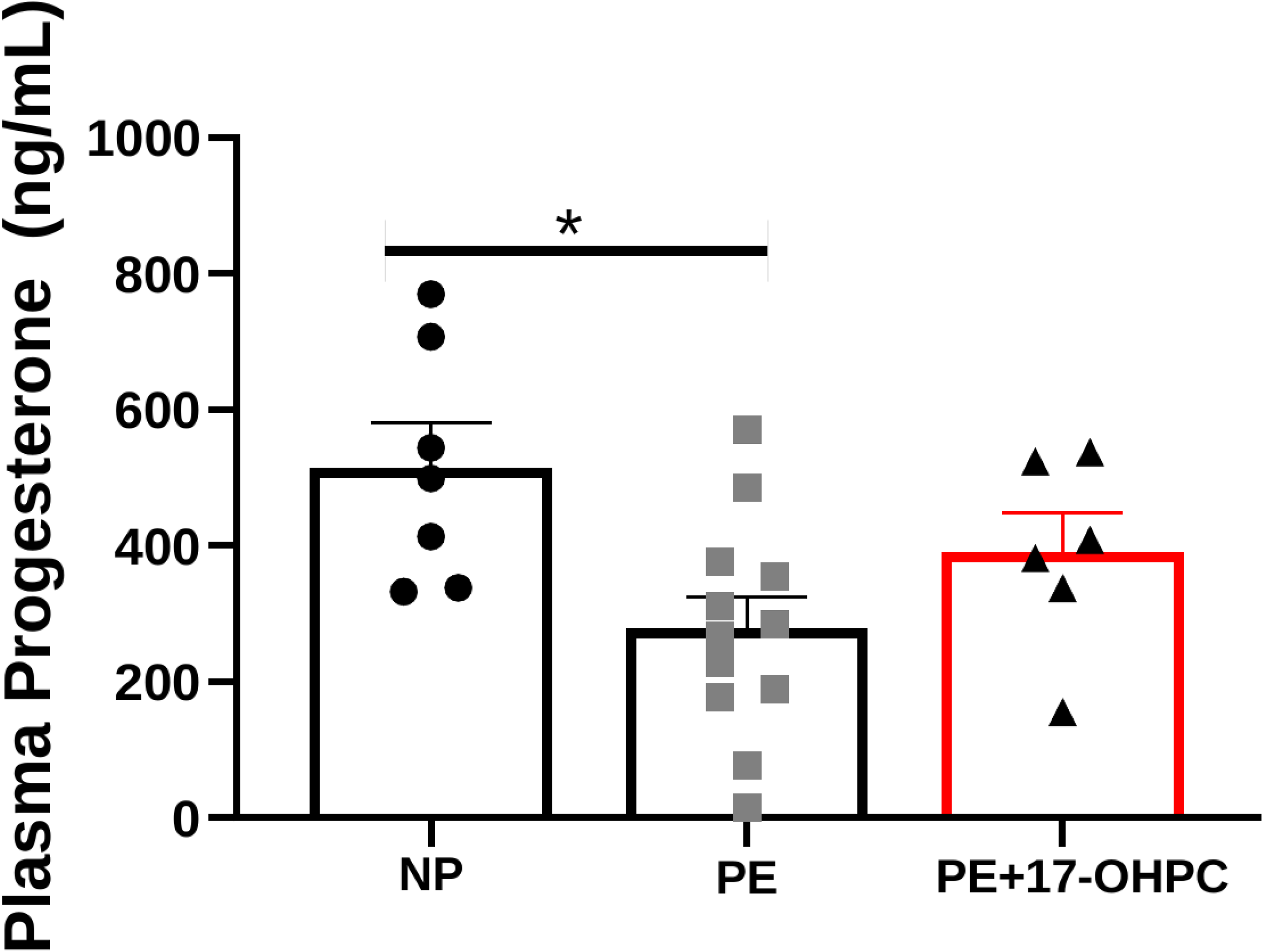
Progesterone levels are reduced in pregnant women with PE. Statistical differences were established using a one-way ANOVA with Holm Sidak multiple comparison post-hoc test. Data are shown as means ± S.E.M. * p< 0.05. * P< 0.05 *vs.* NT and ^#^p< 0.05 *vs.* PE

Although we know progesterone stimulates PIBF, the role for progesterone, in the form of 17**-**OHPC, to stimulate secretion of PIBF in preeclampsia has not been studied and was addressed in this study. Our data showed that pregnant women with PE had lower levels of PIBF and importantly, 17-OHPC supplementation increases circulating PIBF (n=5-6, p<0.05, **Figure 4)**.

**Figure 4.**
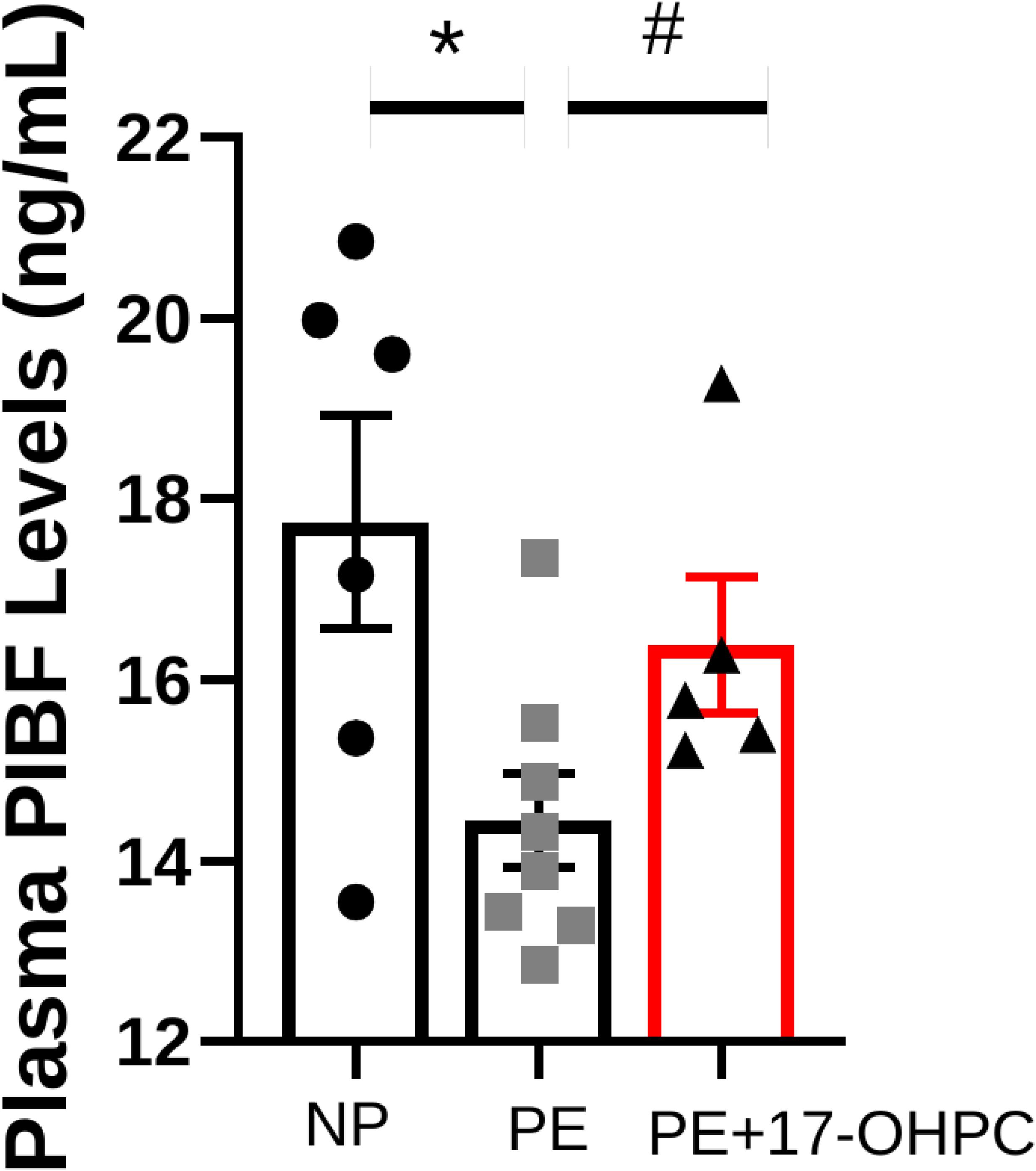
17-OHPC increases PIBF in pregnant women with PE. Statistical differences were established using a one-way ANOVA with Holm Sidak multiple comparison post-hoc test. Data are shown as means ± S.E.M. * p< 0.05. * P< 0.05 *vs.* NT and ^#^p< 0.05 *vs.* PE.

### 17-OHPC reduces Placental PPET-1 in pregnant women with PE

To determine whether levels of ET-1 were higher in pregnant women with PE, we extracted and measured endothelin from plasma. Expression of PPET-1 in placenta samples via RT-PCR. Pregnant women with PE had significantly higher ET-1 compared to pregnant normotensive women (n=4-5, p< 0.05, Figure 5A). Circulating endothelin-1 (ET-1) was 2.53+/− 0.4 pg/mL in NP (n=4), 6.7 +/− 1.4 in PE (n=5, p< 0.05) and 4.9 +/− 1.3 in PE+17-OHPC (n=5). Placental PPET-1 expression was higher in PE pregnancies than in PE+17-OHPC. Placenta ET-1 reduced to 0.96 in PE+17-OHPC (n=4, **Figure 5B**) compared to PE (n=4)

**Figure 5.**
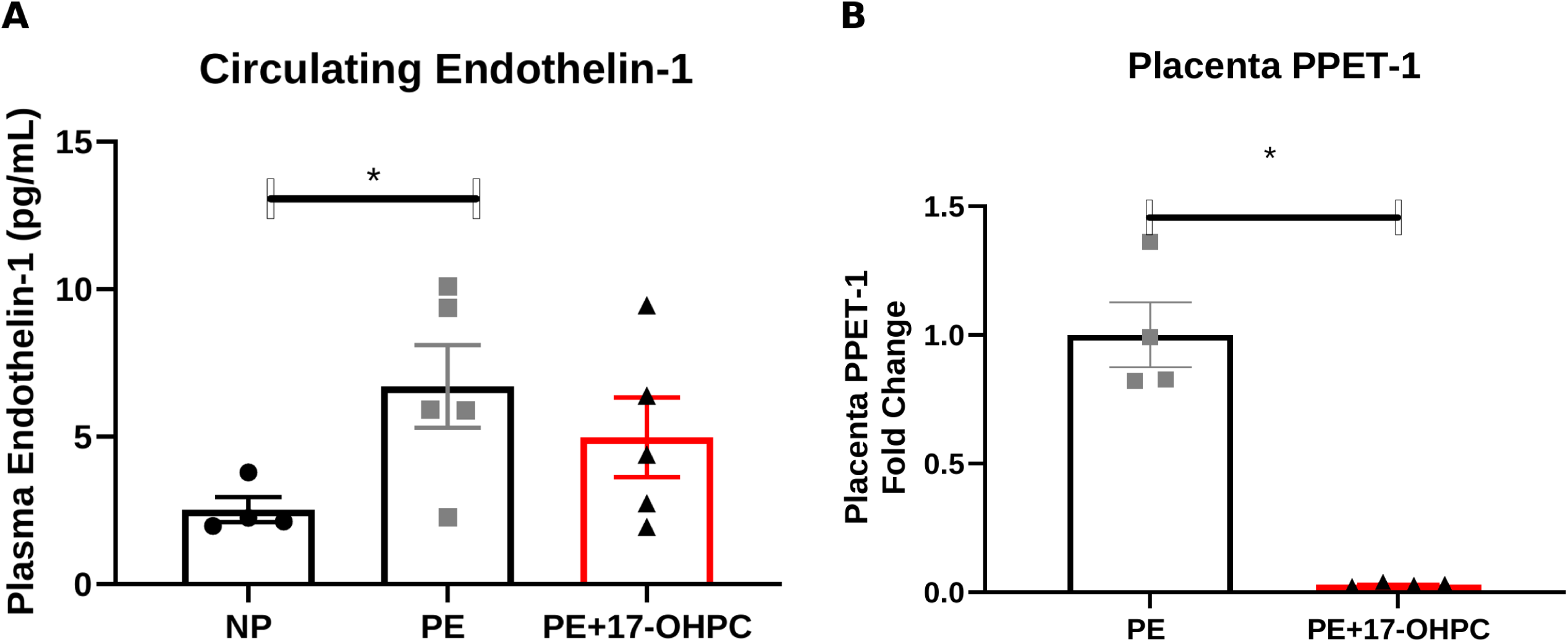
Circulating ET-1 is higher in pregnant women with PE (A). 17-OHPC decreases placental PPET-1 in pregnant women with PE (B). Statistical differences were established using a one-way ANOVA with Holm Sidak multiple comparison post-hoc test (A). Differences between groups and treatments were statistically analyzed using Student t-test (B). Data are shown as means ± S.E.M. * p< 0.05. * P< 0.05 *vs.* NT and ^#^p< 0.05 *vs.* PE.

### 17-OHPC decreases circulating sFlt-1 and increases NO in pregnant women with PE

sFlt-1 was 3742 +/−126 pg/mL in PE group which significantly decreased to 3199+/− 138 in PE+17-OHPC group (n=5-7, p<0.05, **Figure 6A**). Furthermore, nitrate-nitrite levels from plasma were significantly increased after 17-OHPC supplementation. Circulating nitrate-nitrite levels were 50.7±9.3 in PE (n=9), which increased to 94.4±14.2 in PE+17-OHPC group (n=6, p<0.05, **Figure 6B**).

**Figure 6.**
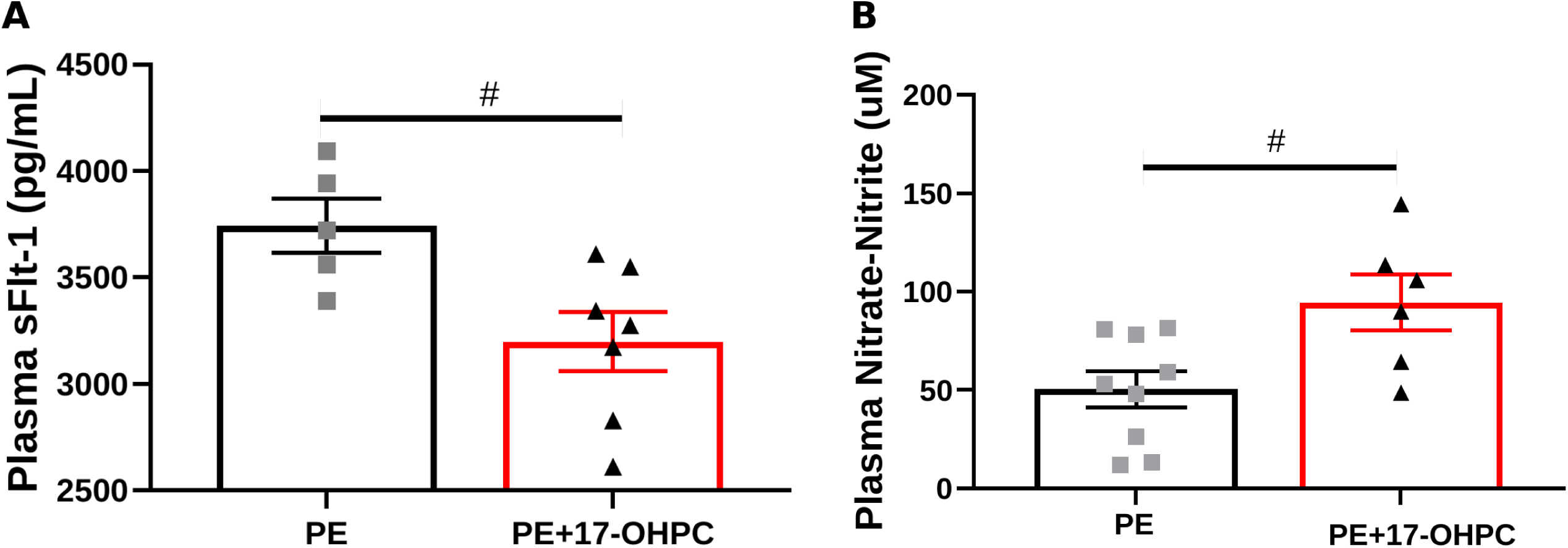
17-OHPC improves circulating sFlt-1 (A) and nitrate-nitrite levels (B) in pregnant women with PE. Statistical differences were established using a one-way ANOVA with Holm Sidak multiple comparison post-hoc test (A). Differences between groups and treatments were statistically analyzed using Student t-test (B). Data are shown as means ± S.E.M. * p< 0.05. * P< 0.05 *vs.* NT and ^#^p< 0.05 *vs.* PE.

### 17-OHPC decreased circulating TNF-α and AT1-AA in pregnant women with PE

TNF-α was 21+/−4 in NP(n=6), 41.3 +/− 3.9 pg/mL in PE (n=8), which decreased to 21.1+/−5.5 in PE+17-OHPC (n=6, p< 0.05, **Figure 7**). AT1-AA were 23+/−5.2 ΔBPM in PE (n=4), which significantly reduced to 10.38+/− 2.1 ΔBPM in PE+17-OHPC (n=8, p< 0.05, **Figure 8)**.

**Figure 7.**
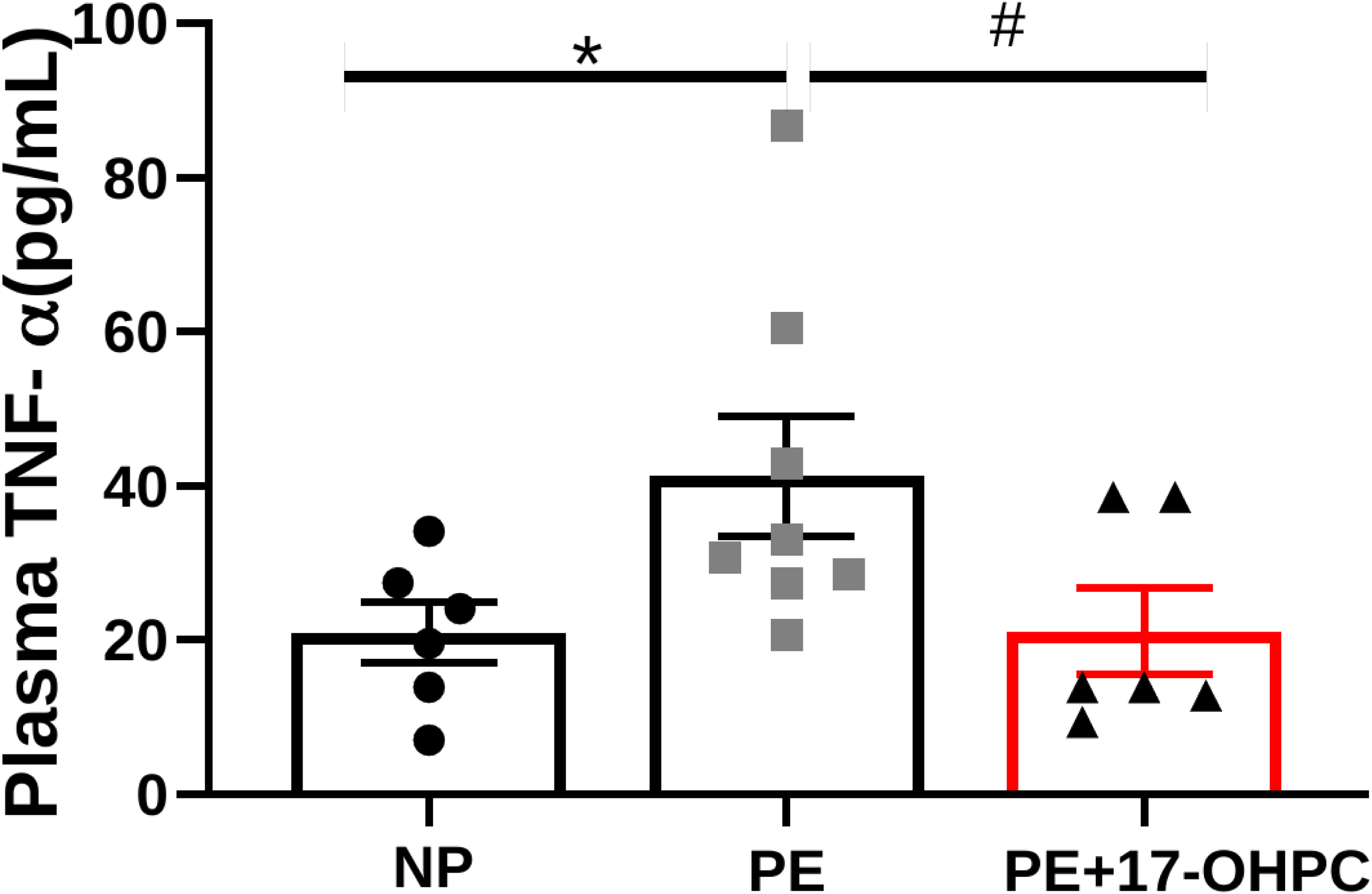
17-OHPC reduces circulating TNF-alpha. Statistical differences were established using a one-way ANOVA with Holm Sidak multiple comparison post-hoc test (A). Data are shown as means ± S.E.M. * p< 0.05. * P< 0.05 *vs.* NT and ^#^p< 0.05 *vs.* PE.

**Figure 8.**
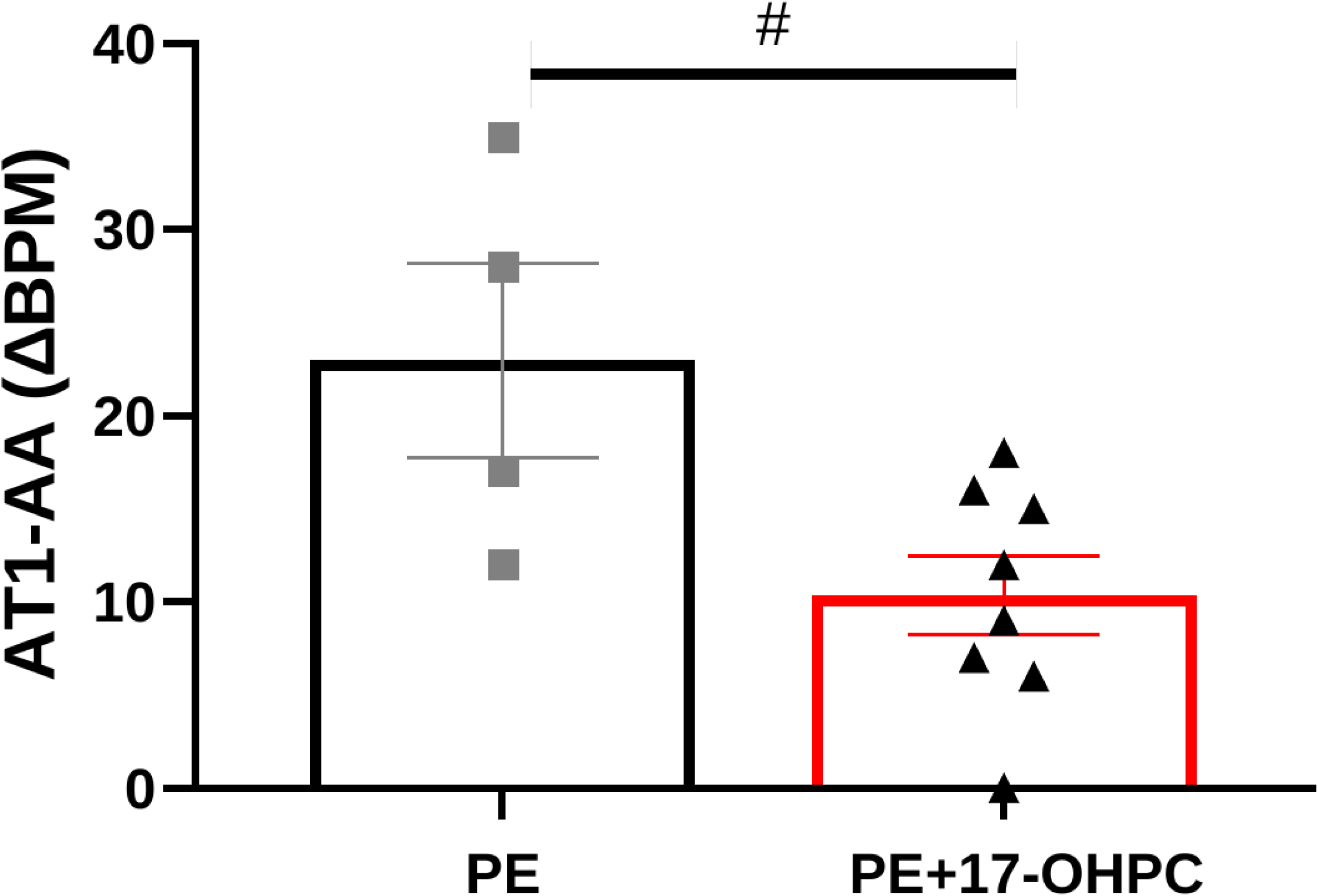
17-OHPC decreases AT1-AA in pregnant women with PE. Statistical differences were established using a student t-test. Data are shown as means ± S.E.M. * p< 0.05. * P< 0.05 *vs.* NT and ^#^p< 0.05 *vs.* PE.

### 17-OHPC reduces placental NK Cells

Placental CD4+ T cells were 2± 0.1% gated cells and NK cells were 6±3 % gated cells in NP controls (n=3), which significantly increased to 6 ±3% (CD4+ T cells) and 21±6 % (NK cells) in the PE group (n=3-4, p<0.03) but reduced to 3 ±1% (CD4+ T cells) and 5 ± 1% gate (NK cells) in the 17-OHPC treatment group (n=3-4,). Although there is an obvious trend toward reduction of T cells, only reduction in NK cells achieved statistical significance in the treatment group.

## Discussion

In this non-randomized open-label clinical trial of intramuscular 17-OHPC in the setting of expectant management of preeclampsia with severe features, we observed a statistically significant decrease in biomarkers of preeclampsia, an increase in NO expression, a decrease in maternal blood pressures, and an increase in latency to delivery in the treatment group with 17-OHPC versus PE controls receiving only the current standard of care.

Many potential factors should be considered in the interpretation our study data. First regarding maternal blood pressure, contrary to the previous clinic trials by Sammour et al., we did not employ progesterone in isolation [17, 24]. In our trial, 17-OHPC was studied as an adjunctive therapy to routine expectant management of preeclampsia with severe features which includes the use of oral and IV antihypertensives. As such, there is obvious confounding from coadministration from other antihypertensive therapies and variation in how aggressively house staff titrated those medicines. However, the trend toward improved blood pressures is consistent with prior published data on progesterone therapy and preeclampsia. In our study design, it was felt to represent an inappropriate risk of harm to patients to withhold other antihypertensive therapies. The benefit of our approach is that it becomes clear that 17-OHPC integrates well into a multi-modal approach of treating preeclampsia and harmonizes well with current standard of care management.

Additionally, it is also worth noting that in the previous data from Sammour et al., the antihypertensive effects of progesterone therapy were found to peak between 6 and 12 days of daily dosing [17, 24]. This trend in therapeutic effect is consistent with the proposed mechanism of action of 17-OHPC in this study. Modification of downstream inflammatory mediators would almost certainly result in a slow temporal trend in blood pressure as has been observed in prior studies. The blood pressure trends noted by Sammour et al. combined with our study findings also help draw a couple significant conclusions. Since the peak antihypertensive effect likely occurs over days, it is unclear how to best capture the antihypertensive changes from 17-OHPC therapy in this study. Our study design and use of adjunctive antihypertensive therapies likely do not allow us to determine the peak antihypertensive effect of 17-OHPC or the exact temporal trend in blood pressure correction that occurs with 17-OHPC administration. Also, the prior data on the subject from Sammour et al. were based on daily administration of natural progesterone. While 17-OHPC has a prolonged mechanism of action versus natural progesterone, it remains unclear if more frequent or higher dose administration of 17-OHPC would result in even better amelioration of maternal blood pressures and other outcomes or if a lower dose of 17-OHPC is non-inferior to the dose we studied. Our study is the only to evaluate treatment of PE with 17-OHPC in humans and the optimal timing and dose of 17-OHPC for treatment of PE has yet to be established.

We noted in our study population, consistent with our prior observations, that patients with EOPE with severe features were progesterone deficient versus normal pregnant controls. The relationship between progesterone deficiency and PE has been discussed in the introduction of this document and has good supporting evidence[17–20]. PIBF deficiency in preeclampsia is less well established in the literature. Importantly, we noted that serum PIBF levels were decreased in PE patients versus NP patients and that PIBF levels increased in PE patients after supplementation with 17-OHPC. This increase in PIBF supports the proposed mechanism that 17-OHPC acts in a PIBF-dependent fashion, and it adds to the biologic plausibility that a correctible decrease in PIBF could be pathophysiologic in PE. It is also possible that PIBF independent of progestogen administration could represent another possible therapeutic strategy for treatment of PE. Furthermore, we found decreased populations of NK cells and levels of downstream inflammatory markers of preeclampsia in the PE patients treated with 17-OHPC versus control PE patients receiving routine management. In the treatment group, TNF-α, sFlt-1, and AT1-AA were reduced versus the PE control group. This corroborates the hypothesis that 17 OHPC acting through PIBF could combat proinflammatory activities occurring in PE patients leading to better blood pressure control and delay to deliver. These decreases in proinflammatory factors led to decreases in sFlt-1 and endothelin, which have been shown in preclinical models to aid in blood pressure lowering effects in response to placental ischemia.

## 4. Limitations and strengths

The limitations of this study are as follows. All human participants were recruited and enrolled at a single center. Neither clinical staff nor patients were formally blinded to treatment groups. Although it is worth noting that enrollment was completed by research staff with minimal involvement from clinicians. Clinical eligibility was reviewed by an MFM fellow to confirm that patients met inclusion and exclusion criteria. Otherwise, the treating physicians were not routinely notified of the assignment of patients to a study arm. Also, patients did not undergo randomization to treatment versus control groups.

17-OHPC injections used in the study which were widely available, and FDA approved for the indication of prevention of preterm birth at study conception. The FDA approval for prevention of recurrent preterm birth was withdrawn during the study period and which drastically affected drug availability and lead the study to close the treatment arm prematurely. However, it is worth noting that this removal of FDA approval for prevention of recurrent preterm birth was not due to maternal to fetal safety outcomes and that our trial did not show an increased in adverse maternal, fetal, or neonatal outcomes in the 17-OHPC treatment group. Placebo injections were not used for the control group. Additionally, enrollment slowed significantly during 2019 and through 2020 related to the COVID pandemic.

## 5. Conclusion

In this novel study of 17-OHCP as an adjunct to standard of care expectant management for treatment EOPE with severe features, we observed a decrease in maternal blood pressures, and a normalization of progesterone and PIBF. We believed this led to lowering of hypertensive mediators, ET-1, sFlt-1 and AT1-AA thereby contributing to an increase in NO expression in PE women versus PE controls receiving only the standard of care. Importantly, there was no increase in adverse outcomes in mothers, fetuses, or neonates seen in patients treated with 17-OHPC.

Further research is needed to establish the optimal progestogen therapy, dosing, and recipient characteristics for patients with PE in larger randomized and blinded trials. Additionally, further research is needed to assess longer term maternal and fetal outcomes and to further establish the mechanisms of progestogens and PIBF in the treatment and pathophysiology of PE.

## Data Availability

The data, analytic methods, and study materials that support the findings of this study will be made publicly available by the corresponding author.

## Notes

### Competing Interest Statement

The authors have declared no competing interest.

### Clinical Trial

ClinicalTrials.gov ID NCT02989025

### Author Declarations

The University of Mississippi Medical Center's IRB approved the study.

